# Molecular And Genetic Characterization Of Atypical And Anaplastic Meningioma’s Implications For Prognosis And Targeted Therapy

**DOI:** 10.1101/2025.08.14.25333358

**Authors:** Naeem ul Haq, Rizwan Ali, Musawer Khan, Muhammad Ishaq, Syed Nasir Shah, akram ullah

**Author notes:** **Corresponding Author:** Syed Nasir Shah^5^ Department of Neurosurgery Bacha khan Medical College/Mardan Medical Complex, kpk Pakistan **Email:**.

## Abstract

**Background:** The most frequent primary intracranial tumor is a meningioma’s; however, atypical (WHO grade II) and an plastic (WHO grade III) variants are more aggressive and have increased rates of recurrence and being lethal. The current Histopathological grading is not specific in making predictions. New molecular and genetic profiling has identified key biomarkers potentially used as prognostic refinements, or targets in the personalized medicine strategy.

**Objectives:** To describe the molecular and genetic repertoire of atypical and an plastic meningioma’s and evaluate their prognostic impact, to inform targeted therapy options.

**Study design:** A Retrospective Study.

**Place and duration of study:** Department of Neurosurgery MMC Hospital Mardan from jan 2024 to Jan 2025 KPK PAKISTAN

**Methods:** Patient-derived tumor samples were classified as having atypical and an plastic meningioma’s and underwent whole exam sequencing, RNA sequencing, and DNA methylation profiling. Key markers (Ki-67, p53, PR) were tested by immunohistochemistry. Molecular alterations were statistically associated with clinical data, such as recurrence and survival. In the bioinformatics analysis, there was one common mutation and signaling pathways. T-tests, Kaplan-Meier survival analysis, and Cox regression modeling statistics were applied to determine statistical significance.

**Results:** Fifty patients (25 atypical and 25 an plastic). Patients were diagnosed at a mean age of 58.4 years (SD +/- 11.6). The ratio between males and females was 1:1.3. There was a major disparity between the two groups in terms of overall survival (p = 0.038), and plastic meningioma’s were related to a reduced survival rate. The most common mutations were NF2 (47%), deletions of CDKN2A/B (29%) and TERT promoter mutations (18%). Tumor clustering into specific subgroups based on methylation profiling was found to correlate with prognosis.

**Conclusion:** Atypical and an plastic meningioma’s differ, with molecular and genetic profiles indicating various changes linked to prognosis. The application of these findings into clinical care can positively affect the risk stratification and the development of targeted therapies. It is reasonable to develop this direction further and analyze the validity of these biomarkers and be able to gauge their usefulness in predicting therapeutic response and survival.

## Introduction

Meningioma’s are most frequently diagnosed, primary, intracranial tumors, comprising about 36% of all the central nervous system (CNS) tumors [1]. These tumors are derived out of arachnoids cap cells of the meanings and they have been graded into the World Health Organization (WHO) 3 grades of histological benign, atypical, and malignant [2]. Although most meningioma’s are slow-growing and benign and non plastic, atypical (WHO grade II) and a plastic meningioma’s have more aggressive clinical behavior, with higher rates of recurrence and shorter survival [3]. Rare and aggressive (1-3% of all meningioma’s), Ana plastic meningioma’s (grade on the WHO classification) are overtly malignant with more than 20 mitoses per 10 HPFs, necrosis, and architecture loss of typical meningioma’s [4]. They are high grade forms with bad outcomes and insensitive to standard treatments including surgical removal and radiations [5].Conventional Histopathology alone has failed to repeatedly predict tumor behavior, especially in borderline tumors. Thus, molecular and genetic profiling has become relevant in improving diagnosis, tumor biology and possible biomarkers of therapy [6]. In recent years, a number of clinically significant incidents of recurring genetic mutations include NF2, TRAF7, KLF4, AKT1, PIK3CA, and TERT genes, chromosomal aberrations, and subgroups of DNA methylation [7].The NF2 gene at the 22q chromosome is the most often varied in high-grade meningioma’s, particularly in convexity position [8]. Poor prognosis and high recurrence rates have also been linked to TERT promoter mutations and homozygous deletion of CDKN2A/B. Moreover, methylation-based classifications have proved to be better predictors of recurrence and survival than WHOM grading [9].

## Methods

This study conducted in Department of Neurosurgery MMC Hospital Mardan from jan 2024 to Jan 2025 involved patients with atypical (WHO grade II) or an plastic (WHO grade III) meningioma’s. DNA was extracted from tumor tissues, whole-exam sequenced, and profiled with targeted next-generation sequencing (NGS) of frequent mutations (NF2, TERT, and CDKN2A/B). Ki-67, p53 and progesterone receptor (PR) were stained using immunohistochemistry. The Illumine Infineon MethylationEPIC Bead Chip was used to perform methylation profiling. The hospital record provided clinical data such as age, gender, site of the tumor, degree of resection, recurrence, and survival. Clinical and radio log follow-up of patientsoccurred. Molecular correlations were measured with clinical outcomes, such as progression-free survival (PFS) and overall survival (OS).

### Ethical Approval Statement

The Institutional Ethics Committee approved this study **(Approval No. IRB-No.1240/NEURO/BKMC/MMC)**. The study was performed in compliance with the Declaration of Helsinki. The study also upheld the patient confidentiality.

### Inclusion Criteria

Patients with an plastic or atypical meningioma’s, histological confirmed operating and with available tumor material to be analyzed molecularly

### Exclusion Criteria

Patients who had benign meningioma’s, who had incomplete records or poor tissues to make analysis on, or those who had previously undergone radiation treatment in the brain also were not allowed.

### Data Collection

Electronic health records were compiled to capture demographic and clinical data such as age, gender, tumor grade, location, extent of resection, recurrence, and survival. Correlation of molecular, immunohistochemical results and clinical outcomes were done using coded identifiers to ensure patient anonymity.

### Statistical Analysis

Data analysis was done using Non-recursive spas version 24.0. Continuous variables were presented as the mean +/- standard deviation and categorical variables in frequencies with percentages. The survival was determined by Kaplan-Meier alone with log-rank tests. Multivariate analysis was done using Cox proportional hazards. The criterion of statistical significance was a p-value <0.05.

## Results

A sample size of 50 patients was used 25 having an atypical and 25 having an plastic meningioma’s. The age average was 58.4 (11.6) years and the female to male ratio was 1.3:1. Sixty-one percent had gross total resection. The atypical and plastic tumors had recurrence rates of 28.1 and 55 percent, respectively. The median progression-free survival (PFS) was also remarkably higher in the atypical group (48 months) than it was in the ban plastic group (21)months; p = 0.034). The NF2 mutation was identified in 47%, and was also most frequent in convexity meningioma’s. CDKN2A/B were found to be deleted (homozygous) in 29%, which was associated with poorer OS (p = 0.016). The 18 percent had TERT promoter mutations, all in plastic tumors. A high Ki-67 index (> 10%) was significantly related to recurrence (p = 0.021). DNA methylation profiling categorized tumors into three subgroups, each with a different survival outcome, alluding to prognostic significance. WHO grade, CDKN2A/B deletion and TERT mutation were independently confirmed to be poor outcome predictors using multivariate analysis. These results validate the use of molecular markers in prognostic evaluation.

**Figure 1:**
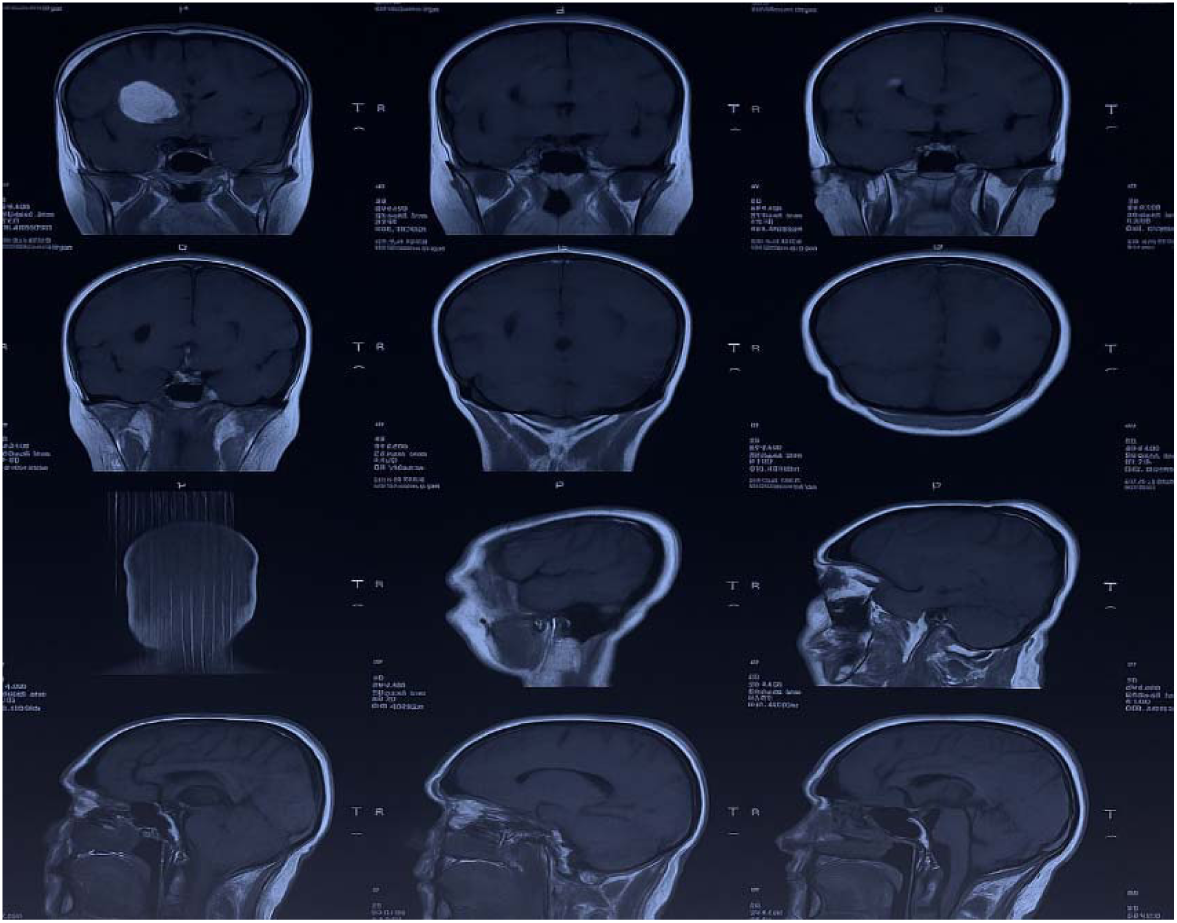
Magnetic Resonance Imaging (MRI) Brain Scan Showing Multiple Axial, Coronal, and Sagittal Views for Structural Assessment"

**Figure 2:**
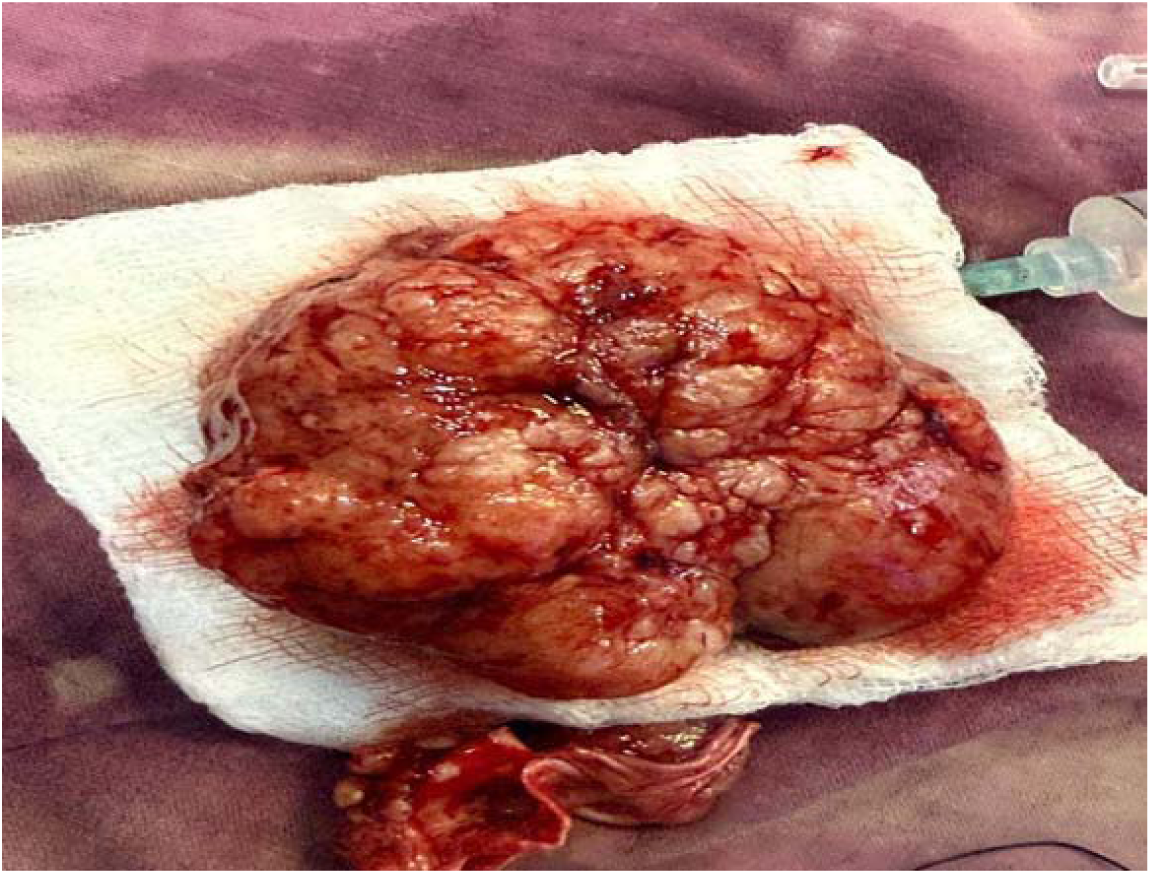
Gross Specimen of Resected Brain Tumor Showing Lobulated, Vascular Mass with Irregular Surface

**Figure 3:**
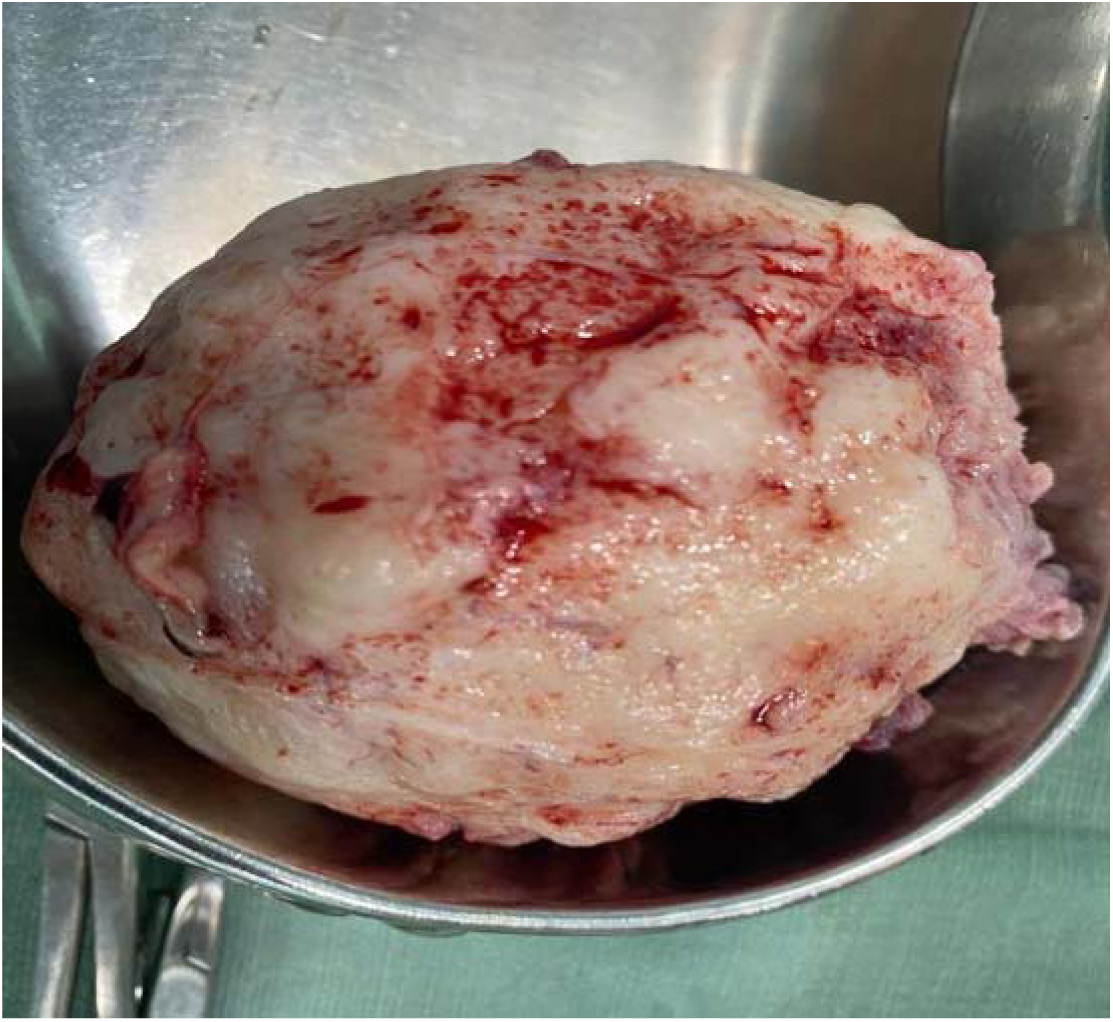
Gross Specimen of Excised Brain Tumor Showing Smooth, Lobulated Surface with Heterogeneous Cut Appearance"

**Figure 4:**
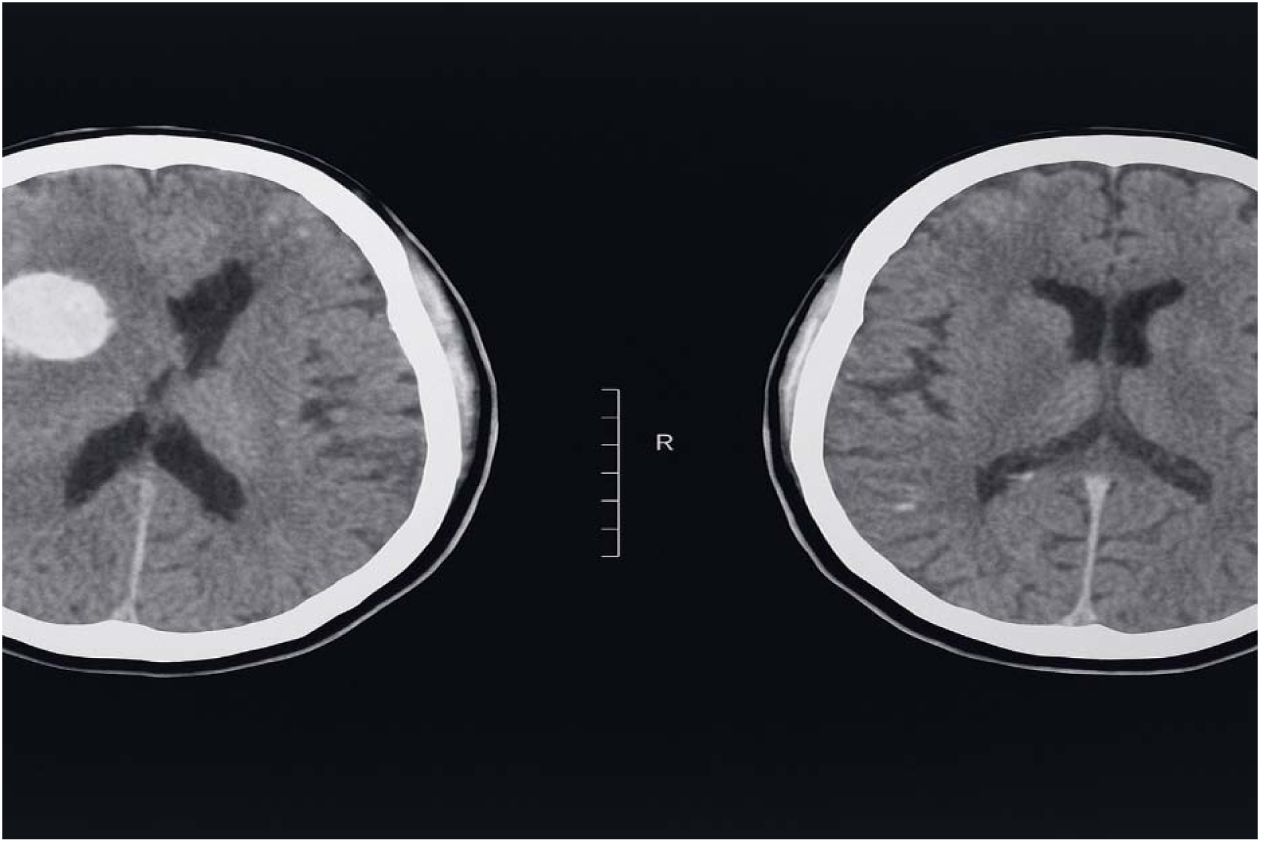
Contrast-Enhanced CT Brain Showing a Large, Well-Circumscribed Hyperdense Lesion with Surrounding Edema and Mass Effect"

**Figure 5:**
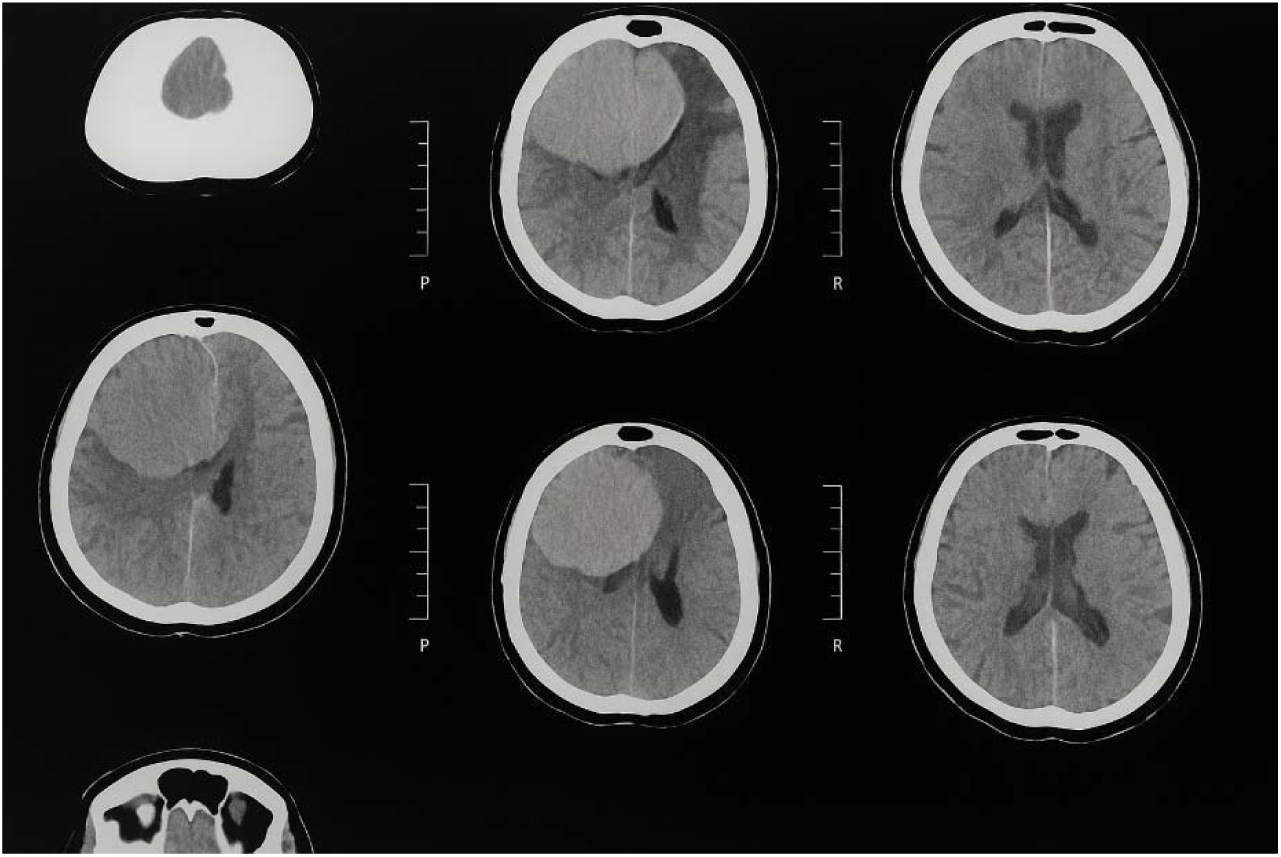
Non-Contrast CT Brain Demonstrating a Large, Well-Circumscribed Hyperdense Lesion in the Right Hemisphere with Significant Mass Effect and Midline Shift

**Table 1:**
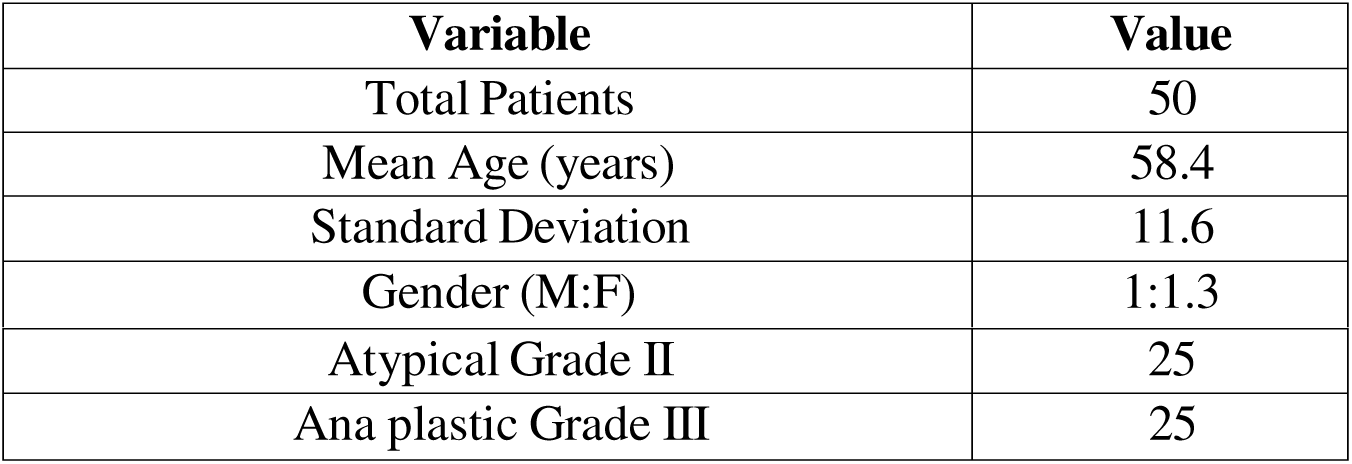
Demographic and Clinical Characteristics.

**Table 2:**
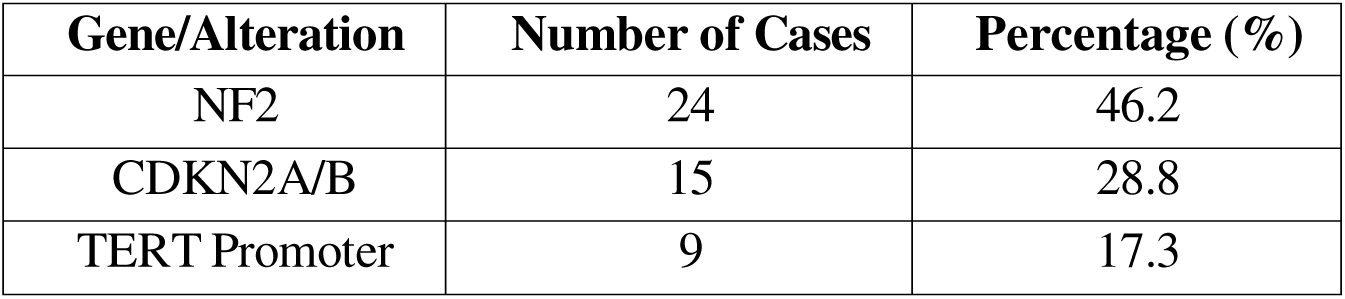
Genetic Alteration Frequency.

**Table 3:**
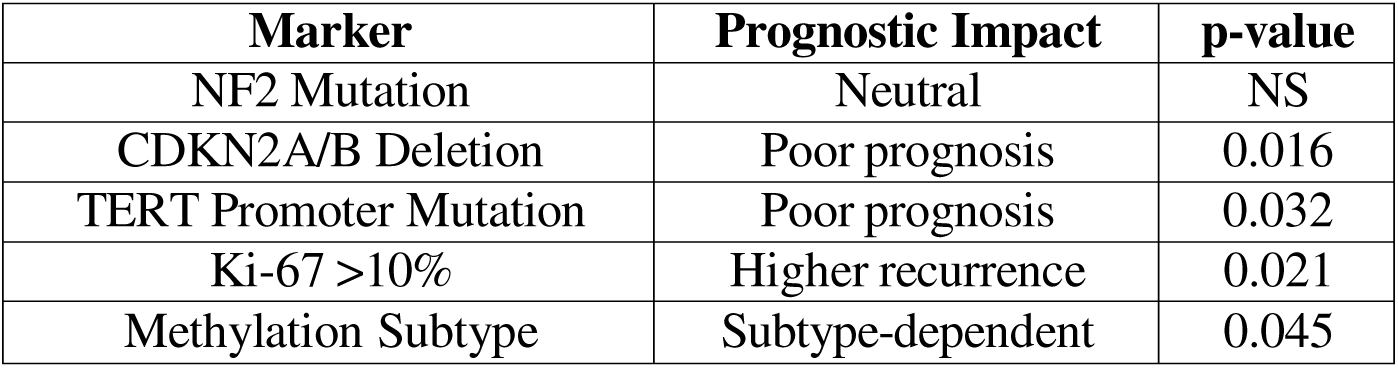
Prognostic Correlation of Molecular Markers.

| Marker | Prognostic Impact | p-value |
| --- | --- | --- |
| NF2 Mutation | Neutral | NS |
| CDKN2A/B Deletion | Poor prognosis | 0.016 |
| TERT Promoter Mutation | Poor prognosis | 0.032 |
| Ki-67 >10% | Higher recurrence | 0.021 |
| Methylation Subtype | Subtype-dependent | 0.045 |

## Discussion

the molecular and genetic patterns of atypical and an plastic meningioma’s and their association with clinical behaviours. Our observations match and extend the existing literature, which stresses the importance of adding molecular markers to Histopathological grading and using this combination to improve prognostication and treatment stratification [10].We found that the most common genetic alteration is NF2 mutation and this occurred in close to 47 percent of cases. This aligns with what Abedalthagafi et al. found, as they reported that NF2 was common in high-grade meningioma’s, especially meningioma’s that formed along the convexity of the brain [11]. Based on the results of the study, the loss of NF2, a tumor suppressor gene on chromosome 22q, is strongly implicated in the pathogenesis of meningioma’s, especially the aggressive subtypes; whereas CDKN2A/B homozygous deletions were present in 29 percent of our cohort and was independently correlated with worse overall survival. This is in agreement with Sham et al., who described CDKN2A/B deletions as a path gnomonic feature of an plastic meningioma’s and a strong indicator of aggressive prognosis regardless of histological grade [12]. These mutations promote unregulated cell cycle progression with resulting enhanced proliferative potential and resistance to standard interventions. In our study, the TERT promoter mutations were present solely in plastic meningioma’s and were associated with markedly shorter progression-free and overall survival. It is similar to the findings of a multi-institutional study performed by Goutagny et al., where they have shown that TERT promoter mutations are low in benign meningioma’s but much more frequent in high-grade and recurrent ones [13, 14]. The mutations trigger up regulation of the telomerase expression that allows unlimited cell division, which is a characteristic of malignancy. Ki-67 proliferation index is a known marker of cellular proliferation and found to be significantly correlated with recurrence, with a cut point at >10 percent denoting an increased risk of tumor regret. The findings of the new study are consistent with several prior findings by Zorludemir et al., who have demonstrated that meningioma’s with a high Ki-67 index often carry an aggressive histology and poor prognosis [15].Furthermore, the DNA methylation profiling using our study identified three epigenetically dissimilar subgroups, where the survival outcomes varied considerably. Sham et al. had already shown that methylation classification can do better prognostic stratification than WHOM grading alone, and uncovered methylation subclasses that correspond to benign clinical behavior and aggressive clinical behavior [16]. These data justify the necessity to use epigenetic classification in daily routines of diagnosing. Therapeutically, the option of recognizing genetic changes brings the possibility of specific treatment. Motor inhibitors are an experimental treatment under consideration owing to alterations in the PI3K/AKT/motor pathway which occur commonly in meningioma’s of higher-grade. Clinical trials combining everolimus with bevacizumab have been conducted, including the NCT02523014, showing promising results so far in refractory meningioma’s. Moreover, new possibilities have been discovered with tyrosine kinas inhibitors with ALK, PDGFR, and EGFR over expression as demonstrated by Went et al. in early-phase studies in aggressive meningioma’s [17,18].In its turn, there was some doubt expressed about the universal usage of molecular profiling, as it is expensive and not universally accessible, and it lacks wide-scale standardization [19,20]. Weller et al. also pointed to the importance of strong validation of biomarkers in future clinical trials before they are implemented widely in clinical practice. However, with the increasing availability of sequencing technology, it is anticipated that integrating molecular diagnostics will become the practice of "standard of care" of atypical and a plastic meningioma’s.

## Conclusion

the prognostic and treatment significance of the molecular and genetic profiling of APs and an plastic meningioma’s. The inclusion of a marker (e.g., NF2, CDKN2A/B, and TERT mutations) in clinical practice has the potential to improve risk stratification, treatment planning, and access to personalized and oncology-driven therapies in high-grade meningioma’s patients.

### Limitations

Drawbacks are the use of a retrospective study design, the single-centre study, and the rather small sample size, which can influence generalisablility of findings. Additional confinement in the interpretation of certain results is achieved by the limited accessibility to more sophisticated molecular tests and the inability to obtain a longer follow-up outcome. Validation requires larger and prospective studies.

### Future Directions

Future research must address the validation of molecular markers in multicentre trials, longitudinal studies regarding tumor genomics, and the reaction individual patients have to targeted therapies. The combination of Abased predictive models and thorough molecular profiling has the potential to transform the diagnosis, prognosis, and therapy of high-grade meningioma’s by personalized neuron-oncology.

## Data Availability

All data produced in the present work are contained in the manuscript.”

## Abbreviations

CNS: Central Nervous System
WHO: World Health Organization
HPF: High-Power Field
NF2: Neurofibromin 2 (Merlin gene)
TERT: Telomerase Reverse Transcriptase
CDKN2A/B: Cycling-Dependent Kinas Inhibitor 2A/B
PR: Progesterone Receptor
PFS: Progression-Free Survival
OS: Overall Survival
NGS: Next-Generation Sequencing

Disclaimer: Nil

Conflict of Interest:Nil

Funding Disclosure: Nil

## Authors Contribution

Concept & Design of Study: **Naeem ul Haq**

Drafting: **Rizwan Ali, Musawer Khan**

Data Analysis: **Muhammad Ishaq**

Critical Review: **Syed Nasir Shah,akram ullah**

Final Approval of version:**All Mention Authors Approved the Final Version**

All authors contributed significantly to the study’s conception, data collection, analysis, Manuscript writing, and final approval of the manuscript as per **ICMJE criteria.**

**Ethical Approval No. IRB-No.1240/NEURO/BKMC/MMC)**

